# Multi-center evaluation of baseline neutrophil:lymphocyte (NLR) ratio as an independent predictor of mortality and clinical risk stratifier in Idiopathic Pulmonary Fibrosis

**DOI:** 10.1101/2022.04.29.22274470

**Authors:** Theresia A Mikolasch, Peter M. George, Jagdeep Sahota, Thomas Nancarrow, Shaney L Barratt, Felix A. Woodhead, Vasilis Kouranos, Victoria S A Cope, Andrew W Creamer, Silan Fidan, Balaji Ganeshan, Luke Hoy, John A Mackintosh, Robert Shortman, Anna Duckworth, Janet Fallon, Helen Garthwaite, Melissa Heightman, Huzaifa I Adamali, Sarah Lines, Thida Win, Rebecca Wollerton, Elisabetta A Renzoni, Matthew Steward, Athol U. Wells, Michael Gibbons, Ashley M Groves, Bibek Gooptu, Chris J. Scotton, Joanna C. Porter

**Affiliations:** CITR, UCL Respiratory, UCL, London; Interstitial Lung Disease Service, UCLH NHS Trust, London; Interstitial Lung Disease Unit, Royal Brompton Hospital; National Heart and Lung Institute, Imperial College London; College of Medicine & Health, University of Exeter; Academic Department of Respiratory Medicine, Royal Devon & Exeter NHS Foundation Trust; Bristol Interstitial Lung Disease Service, North Bristol NHS Trust; Academic Respiratory Unit, University of Bristol; Institute for Lung Health and Leicester Interstitial Lung Disease Service and NIHR Leicester Biomedical Research Centre - Respiratory, Glenfield Hospital, Groby Road, Leicester; Department of Respiratory Sciences and Leicester Institute of Structural & Chemical Biology University of Leicester; Institute of Nuclear Medicine, UCL and Department of Nuclear Medicine UCLH; The Prince Charles Hospital, Queensland, Australia; Department of Respiratory Medicine, Somerset Lung Centre, Musgrove Park Hospital, Taunton; Lister Hospital, North East Herts Trust, Stevenage

## Abstract

**Background:** Idiopathic pulmonary fibrosis (IPF) is a progressive, fatal disorder with a variable disease trajectory. The aim of this study was to assess the potential of neutrophil-to-lymphocyte ratio (NLR) to predict outcomes for people with IPF.

**Method:** We adopted a two-stage discovery and validation design using patients from the UCL partners (UCLp) cohort. For the discovery analysis, we included 71 patients from UCLH. In the validation analysis, we included 928 people with IPF, using real-life data from UCLH and 5 other UK centres. Data were collected from patients presenting over a 13-year period with a mean follow up time of 3.7 years.

**Findings:** In the discovery analysis, we showed that values of NLR (<2.9 vs >/=2.9) were associated with increased risk of mortality (HR 2.04, 95% CI 1.09-3.81; p=0.025). In the validation cohort we confirmed this association of high NLR with mortality (HR 1.65, 95% CI 1.39-1.95; p<0·0001) and showed incorporation of baseline NLR in a modified GAP-stage/index (GAP/index)-plus improved predictive ability

**Interpretation:** We have identified NLR as a widely available test that significantly correlates with lung function, can predict outcomes in IPF and refines clinical GAP-staging. NLR may help ILD specialist centres prioritise at risk patients in a timely way, even in the absence of lung function.

## Introduction

Idiopathic pulmonary fibrosis (IPF) is a progressive, fatal disorder with a very variable disease trajectory. Available treatments for IPF are expensive and merely slow disease progression with frequent side-effects. A prognostic biomarker would guide treatment decisions, timing of lung transplant or end of life care and help patients and clinicians to plan.

Clinical cohort staging in IPF relies on the Gender, Age, Physiology (GAP) index (a score from 0 to 7) with associated GAP stage (I-III), a static measure unable to identify rapidly deteriorating patients, or assess treatment response. There is an unmet need for biomarkers to guide a personalized approach to care, as well as for cohort stratification in clinical trials. Only two biomarkers have been validated to refine the GAP staging system by identifying high and low risk patients within a given GAP stage. The first used a 52-gene expression signature, an approach that requires calibration against a control cohort ^6^, and the second measured glucose uptake in the lung with Positron Emission Tomography (PET) ^7^. Both biomarkers require specialist expertise and are costly, limiting their practicality. The ideal biomarker would be measurable in the blood using a simple and widely available test and would predict prognosis and potentially response to therapy.

The Neutrophil-Lymphocyte Ratio (NLR) is easily and inexpensively measured from a complete blood count (CBC), and has indicated severity in studies of diabetes, ^8,9^ cardiovascular ^10^and renal disease ^11^, COPD ^8^, malignancy ^12^and COVID-19 ^13,14^. NLR can also predict development and severity of lung fibrosis in patients with systemic sclerosis^15^, dermatomyositis/polymyositis ^16^ and a composite endpoint of ‘absolute decline in 6MWD ≥50 m or death’ at 12 months in IPF ^4^. It is not known whether addition of NLR to GAP will refine clinical cohort staging in IPF and guide management.

Here we use a two-stage derivation and validation model to determine the potential of NLR to refine the predictive prognostic power of the clinical GAP score in IPF

## Methods

### Study design and participants

An observational study to evaluate NLR as an independent mortality risk predictor in IPF. Derivation cohort, 71 IPF patients enrolled at UCLH (2008-18). Validation cohort of 928 patients comprising: Internal validation cohort, 134 patients from UCLH (2006-18; internal validation cohort); External validation, 794: 279 IPF patients from Royal Brompton and Harefield NHS Trust (RBH) (2006-2018); combined cohort of 515 IPF patients from The Royal Devon and Exeter (RDE) Hospital (300), North Bristol (NB) NHS Trust (85), Taunton and Somerset (TS) NHS Foundation Trust (30); and University Hospitals of Leicester (UHL; 90), (2011-2019). Total cohort size for derivation and validation, n=999. Inclusion criteria: diagnosis of IPF; baseline pulmonary function tests and CBC. Exclusions: malignancy or haematological disorder; infection at time of CBC (CRP >/=20 g/L, clinical/imaging signs of infection); cytotoxic drugs. Exclusion from derivation cohort if on prednisolone >/= 5mg or equivalent at time of CBC, or if insufficient data. CBC was taken at time of diagnosis and analysed locally.

We have previously reported our initial UCLH internal derivation and validation cohorts, 118 patients, as an abstract ^17^. The 515 patients from NB/RDE/TS/UHL were reported as part of a larger cohort comparing basic outcome predictors in IPF versus fHP ^5^

### Outcomes

Primary outcome measure was transplant free survival from CBC measurement to death (all-causes) or transplant in high and low NLR groups using the following censoring: UCLH 28/6/2018, RBH 30/1/2020, RDE combined cohort 12/7/2019. Secondary outcome was assessment of NLR as a mortality predictor independent of GAP index.

### Statistical analysis

A non-biased empirical Cumulative Distribution Function, eCDF plot of baseline NLR of the derivation cohort was used to determine the median NLR. Harrell’s concordance (C)-index was used to determine the ability of NLR to predict outcome accurately with increasing time from baseline. Analysis was performed using STATA 15 (Stata Corp, College Station, Texas). Fisher’s exact test and unpaired two tailed t-tests were used to calculate significance between different group characteristics. All p-values are reported for two-sided confidence intervals. A p-value of <0.05 was considered significant.

### Survival analysis

Both transplant and death were events. Univariate analysis was used to calculate risk of death/prediction of transplant-free survival and the relationship between NLR, NLR category (high/low), GAP Index, GAP Stage, age, sex, FVC (% predicted), TLco (% predicted), steroid therapy (as a binary variable), and transplant-free survival. Significance testing between groups on Kaplan-Meier curves was performed using non-parametric log-rank test. Multivariate stepwise forward cox proportional hazards regression was used to determine whether NLR (as a continuous parameter or category) was independent of the GAP index/stage (and their individual components) and steroids in predicting patient transplant-free survival.

### GAP-Plus and GAP Index Plus

GAP Index Plus was GAP Index plus one additional point added for a high NLR reading GAP-Plus was constructed by up-staging patients’ GAP stage by one if they were in the high NLR category. Thereby creating a four category GAP stage such that GAP Index of 0-3 = Stage 1; GAP Index of 0-3 plus high NLR = Stage 2; GAP Index of 4-5 = Stage 2; GAP Index of 4-5 plus high NLR = Stage 3; GAP Index of 6-8 = Stage 3; GAP Index of 6-8 plus high NLR = Stage 4

### Ethical approvals

Ethical approval was granted by the HRA and Health and Care Research Wales (HCRW). (REC reference: 18/LO/0937). Site specific and local R&D approvals were obtained at each participating site.

## Results

Patient characteristics in the individual and pooled cohorts are summarised in Table 1. For the 999 patients in the combined (discovery and validation) cohorts, there were 533 events (death or transplant) recorded.

**Table 1:**
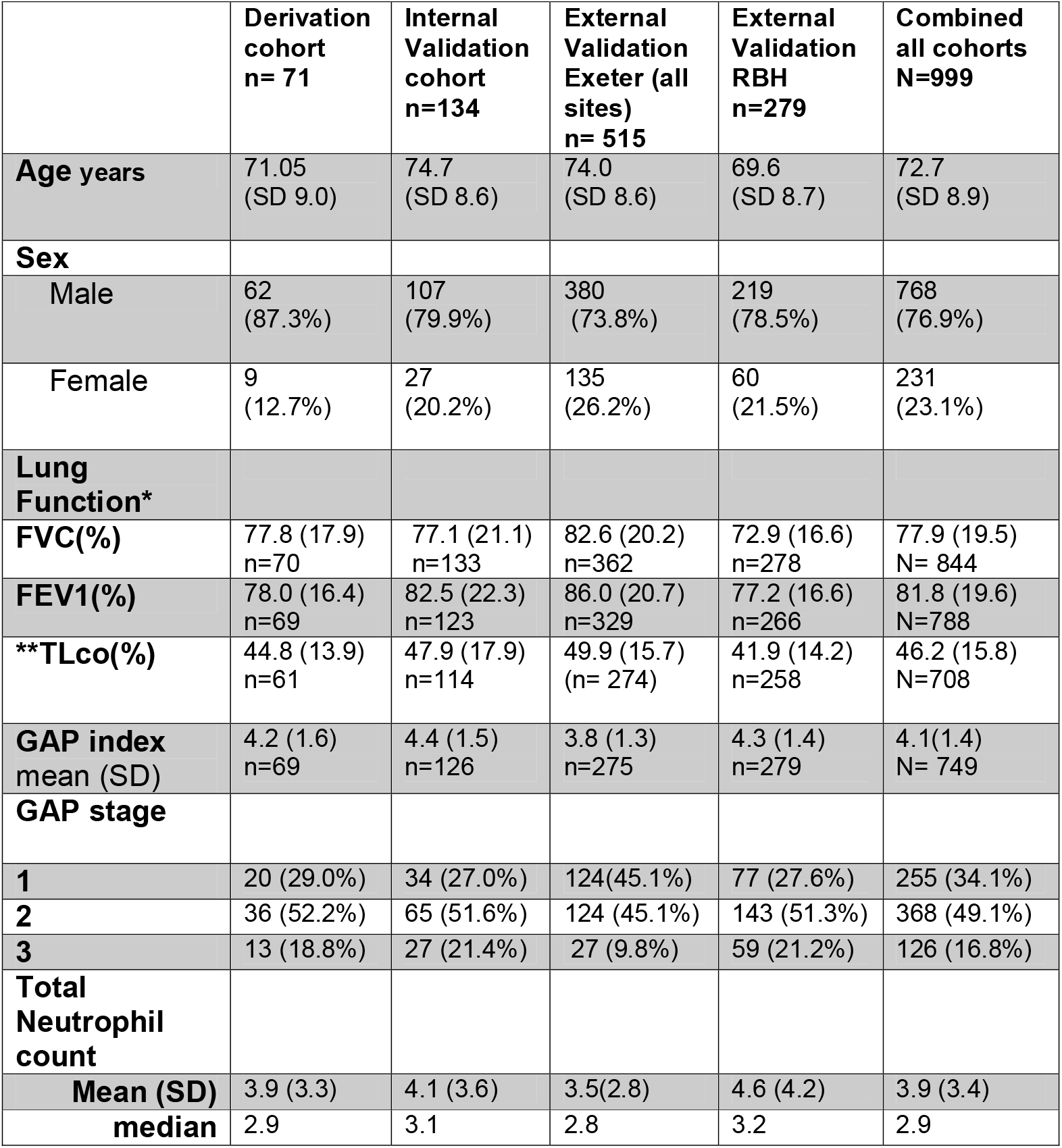
Patient characteristics in derivation and validation cohorts. * Lung Function was taken where available. In some cases, FVC was recorded without FEV1 (the latter is not part of GAP score). **TLco : missing values were only included in GAPscore if documented as ‘not able to perform’-scoring 3 points. Not performed or not recorded scored 0.

|  | **Derivation cohort**<br>**n= 71** | **Internal Validation cohort**<br>**n=134** | **External Validation Exeter (all sites)**<br>**n= 515** | **External Validation RBH**<br>**n=279** | **Combined all cohorts**<br>**N=999** |
| --- | --- | --- | --- | --- | --- |
| **Age years** | 71.05<br>(SD 9.0) | 74.7<br>(SD 8.6) | 74.0<br>(SD 8.6) | 69.6<br>(SD 8.7) | 72.7<br>(SD 8.9) |
| **Sex** |  |  |  |  |  |
| Male | 62<br>(87.3%) | 107<br>(79.9%) | 380<br>(73.8%) | 219<br>(78.5%) | 768<br>(76.9%) |
| Female | 9<br>(12.7%) | 27<br>(20.2%) | 135<br>(26.2%) | 60<br>(21.5%) | 231<br>(23.1%) |
| **Lung Function\*** |  |  |  |  |  |
| **FVC(%)** | 77.8 (17.9)<br>n=70 | 77.1 (21.1)<br>n=133 | 82.6 (20.2)<br>n=362 | 72.9 (16.6)<br>n=278 | 77.9 (19.5)<br>N= 844 |
| **FEV1(%)** | 78.0 (16.4)<br>n=69 | 82.5 (22.3)<br>n=123 | 86.0 (20.7)<br>n=329 | 77.2 (16.6)<br>n=266 | 81.8 (19.6)<br>N=788 |
| **\*\*TLco(%)** | 44.8 (13.9)<br>n=61 | 47.9 (17.9)<br>n=114 | 49.9 (15.7)<br>(n= 274) | 41.9 (14.2)<br>n=258 | 46.2 (15.8)<br>N=708 |
| **GAP index mean (SD)** | 4.2 (1.6)<br>n=69 | 4.4 (1.5)<br>n=126 | 3.8 (1.3)<br>n=275 | 4.3 (1.4)<br>n=279 | 4.1(1.4)<br>N= 749 |
| **GAP stage** |  |  |  |  |  |
| **1** | 20 (29.0%) | 34 (27.0%) | 124(45.1%) | 77 (27.6%) | 255 (34.1%) |
| **2** | 36 (52.2%) | 65 (51.6%) | 124 (45.1%) | 143 (51.3%) | 368 (49.1%) |
| **3** | 13 (18.8%) | 27 (21.4%) | 27 (9.8%) | 59 (21.2%) | 126 (16.8%) |
| **Total Neutrophil count** |  |  |  |  |  |
| **Mean (SD)** | 3.9 (3.3) | 4.1 (3.6) | 3.5(2.8) | 4.6 (4.2) | 3.9 (3.4) |
| **median** | 2.9 | 3.1 | 2.8 | 3.2 | 2.9 |

The median NLR in the derivation cohort was 2.9 and using this cut-off to determine low (>/=2.9) or low NLR (<2.9). Increasing age, male sex, and worse lung function parameters were all associated with the high NLR category (Table 2).

**Table 2:** Baseline and clinical characteristics of the patients (combined cohort, n=999) in low (<2.9, n=502) and high (>/=2.9, n=497) NLR risk groups * p values were calculated by unpaired two-tailed t-test except for sex and GAP stage where a Fisher’s exact test was used.

|  | Low NLR<br>n=502 | High NLR<br>n=497 | p value* |
| --- | --- | --- | --- |
| **Age years (SD)** | 71.9 (9.0)<br>95% CI (71.1-72.6) | 73.5 (8.7)<br>95% CI (72.7-74.3) | **0.0054** |
| **Sex** |  |  | **0.009** |
| **Female** | 134 (26.7%) | 97 (19.5%) |  |
| **Male** | 368 (73.3%) | 400 (80.5%) |  |
| **Spirometry** |  |  |  |
| **FVC(%)** | 80.2 (SD 20.2)<br>95% CI (78.3-82.1) | 75.5 (SD 18.5)<br>95% CI (73.7-77.3) | **0.0004** |
| **FEV1(%)** | 83.4 (19.7)<br>95% CI (81.5-85.4) | 80.1 (19.3)<br>95% CI (78.2-82.0) | **0.016** |
| **TLco % predicted (SD) (n=707)** | 48.8 (15.4)<br>95% CI (47.2-50.4) | 43.5 (15.7)<br>95% CI (41.9-45.2) | **<0.0001** |
| **GAP index** | 3.9 (1.4)<br>95% CI (3.7-4.0) | 4.4 (1.4)<br>95% CI (4.2-4.5) | **<0.0001** |
| **GAP stage** | N=372 | N=377 | **<0.0001** |
| **1** | 154 (41.4%) | 101 (26.8%) |  |
| **2** | 170 (45.7%) | 198 (52.5%) |  |
| **3** | 48 (12.9%) | 78 (20.7%) |  |

When the patients were taken as a combined cohort of 999 and were divided into high NLR (>/=2.9) or low NLR (<2.9) at time 0; there was a significant difference in the median survival between high and low NLR groups (Figure 1; p <0.0001). Median survival in the low NLR group (n=502) of 49.8 months (IQR 24.8-88.3), incident rate of 0.013 and a total time at risk of 17707 months; median survival in the high NLR group (n=497) of 35.9 months (IQR 15.1-63.7, incident rate of 0.021 and a total time at risk of 14426 months (Figure 1).

**Figure 1:**
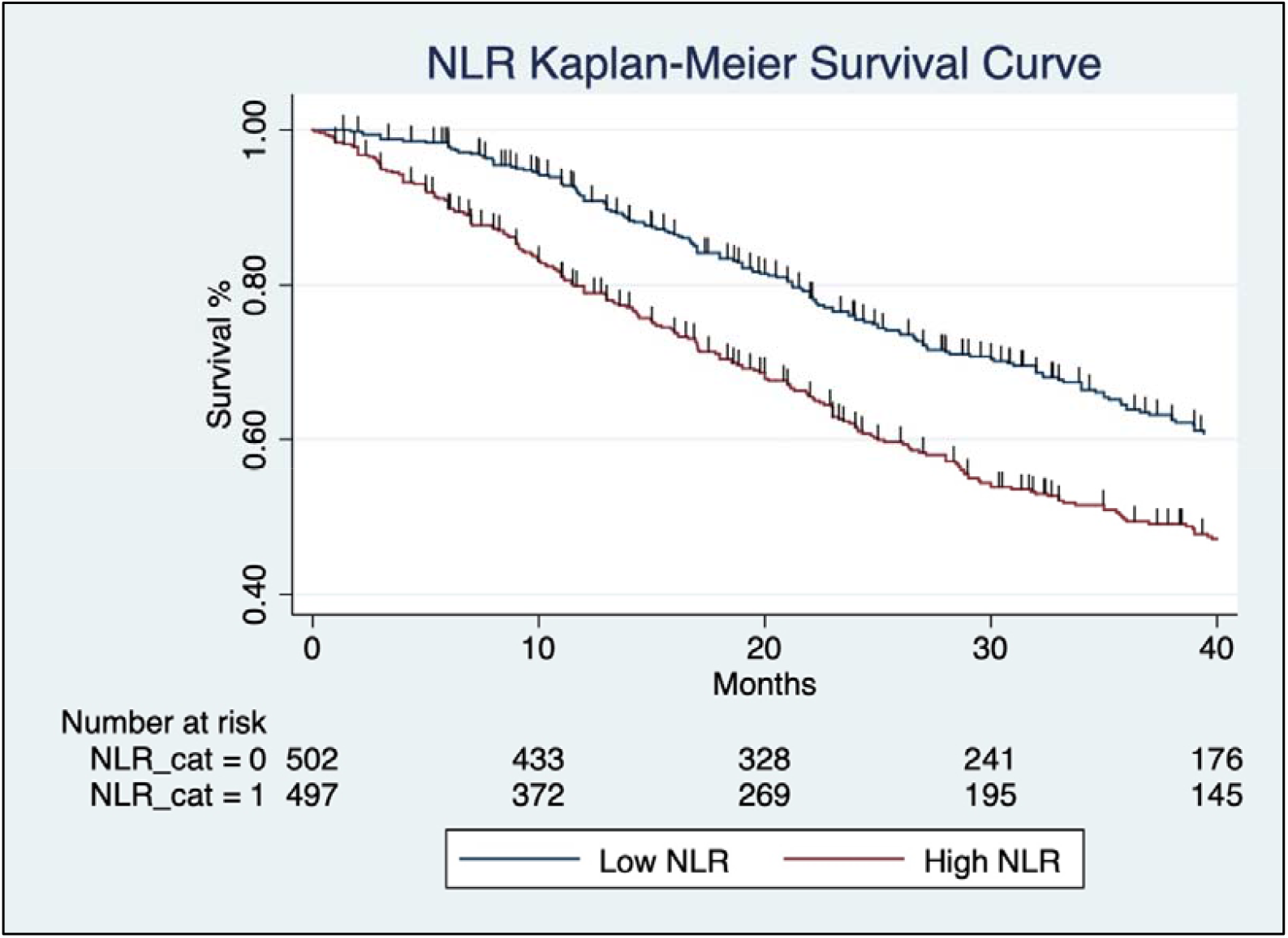
Kaplan-Meier survival curve for all-cause mortality following a diagnosis of IPF for patients in low (<2.9, n=502) and high (>/=2.9, n=497) NLR category at baseline, with follow up of 40 months. The numbers of patients at risk at 10,20,30,40 months for each of these groups is shown in the table immediately below the survival curves This demonstrates a significant difference in mortality between high and low categories (log rank test, p< 0.0001)

In the combined cohort the association of NLR category with GAP stage or GAP Index was highly significant (Table 2; p <0.0001). Although gas transfer data was only available for 71% of subjects, a lower TLco was significantly associated with high NLR (42.2% pred. versus 47.7%, p<0.0001).

Observed mortality was similar to that predicted by GAP stage in the combined patient cohort of 999 patients (Supplementary Table 1). In the pooled validation cohort of 928 patients, the median survival was 44.6 months (interquartile range, IQR, 21-77.6 months) with an incident rate of 0.17/ month and a total time at risk of 29822 months. Therefore, a total of 2485 patient years were analysed.

Median survival per GAP stage is summarised in Table 3. Median survival as stratified by NLR risk category was not significantly different for GAP stage 1 (p =0.245) or 3 (p=0.1381) but was significant for GAP stage 2 (p=0.0127) and for the remaining patients who had no GAP stage recorded due to insufficient lung function data. (p=0.0015; Table 4).

**Table 3:** Median survival per GAP stage for combined cohort (n=999 of which GAP not recorded in n=250)

| **GAP Stage**<br>**n=749** | **Incidence rate** | **Median survival in months (IQR)** |
| --- | --- | --- |
| **GAP 1**<br>**n= 255** | 0.010 | 73.7 (36.0-100.9) |
| **GAP 2**<br>**n= 368** | 0.020 | 40.7 (19.5-60.0) |
| **GAP 3**<br>**n= 126** | 0.037 | 18.0 (10.0-33) |
| **GAP unrecorded**<br>**n= 250** | 0.013 | 60 (22-120.6) |

**Table 4:** Median survival per GAP stage stratified for low (<2.9) and high (>/=2.9) NLR category, n=999 of which n=250 had unrecorded GAP. *2-sided Fisher’s exact

| **GAP Stage and NLR category** | **Incidence rate** | **Median survival months** | **Recorded Failures** | **p- value\*** |
| --- | --- | --- | --- | --- |
| **GAP 1**<br>**NLR low**<br>**n= 154** | 0.009 | 76.1 (41.8-100.9) | 61 |  |
| **GAP 1**<br>**NLR high**<br>**n= 101** | 0.011 | 55.8 (33.2-92.5) | 48 | .245 |
| **GAP 2**<br>**NLR low**<br>**n= 170** | 0.017 | 44 (22.3-65.0) | 94 |  |
| **GAP 2**<br>**NLR high**<br>**n= 198** | 0.023 | 35.7 (16.6-54.2) | 137 | **0.0127** |
| **GAP 3**<br>**NLR low**<br>**n= 48** | 0.031 | 19.5 (13.0-39.4) | 41 |  |
| **GAP 3**<br>**NLR high**<br>**n= 78** | 0.042 | 16.9 (8.2-33) | 67 | .1381 |
| **GAP unrecorded**<br>**NLR low**<br>**n= 130** | 0.009 | 83 (34-120.6) | 39 |  |
| **GAP unrecorded**<br>**NLR high**<br>**n= 120** | 0.018 | 44 (12-74) | 51 | **.0015** |

The difference in expected versus observed events for patients in high and low NLR categories was significant with 235 observed events out of 300.95 expected in the low NLR group versus 303 observed events out of 237.05 expected in the high NLR group in the combined cohort (log rank test, p< 0.0001; Figure 1). Differences between survival in patients with different GAP stages 1-3 (Figure 2A) and GAP Index scores (not shown) reached significance (log rank test, p< 0.0001). Stratifying patients in the same GAP stage by NLR category (low/high) also showed significant differences in survival between low and high NLR for each stage (log rank test, p< 0.0001; Figure 2B-D). The survival differences between the GAP-Plus groups of patients also reached significance (HR 1.80, 95% CI 1.60-1.98; log rank test, p< 0.0001; Figure 3).

**Figure 2:**
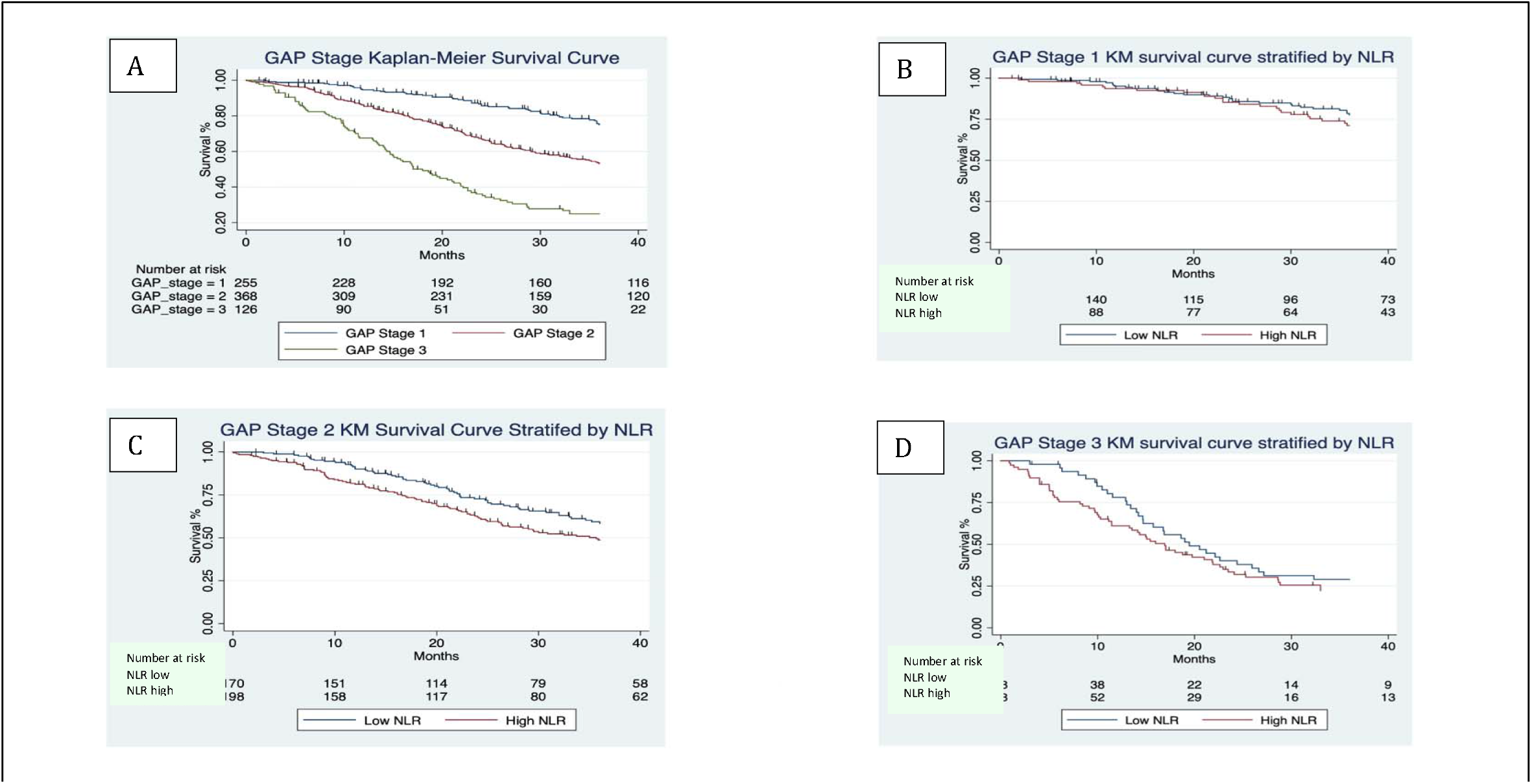
Kaplan-Meier survival curves for all-cause mortality following a diagnosis of IPF with follow up extending to 40 months: A, All patients in combined cohort (n=999) divided into GAP stages 1 (n=255), 2 (n=368) and 3 (n=136); B, Patients in GAP Stage 1 stratified into low (<2.9, n=154) and high (>/=2.9, n=101) NLR category at baseline; C, Patients in GAP Stage 2 stratified into low (<2.9, n=170) and high (>/=2.9, n=198) NLR category at baseline; D, Patients in GAP Stage 3 stratified into low (<2.9, n=48) and high (>/=2.9, n=78) NLR category at baseline. The numbers of patients at risk at 10,20,30,40 months for each of these groups is shown in the table immediately below the survival curves. Differences between survival in patients with different GAP stages 1-3 (Figure 3A) reached significance (log rank test, p< 0.0001). Stratifying patients in the same GAP stage by NLR category (low/high) also showed significant differences in survival between low and high NLR for each stage (log rank test, p< 0.0001; Figure 3B-D).

**Figure 3:**
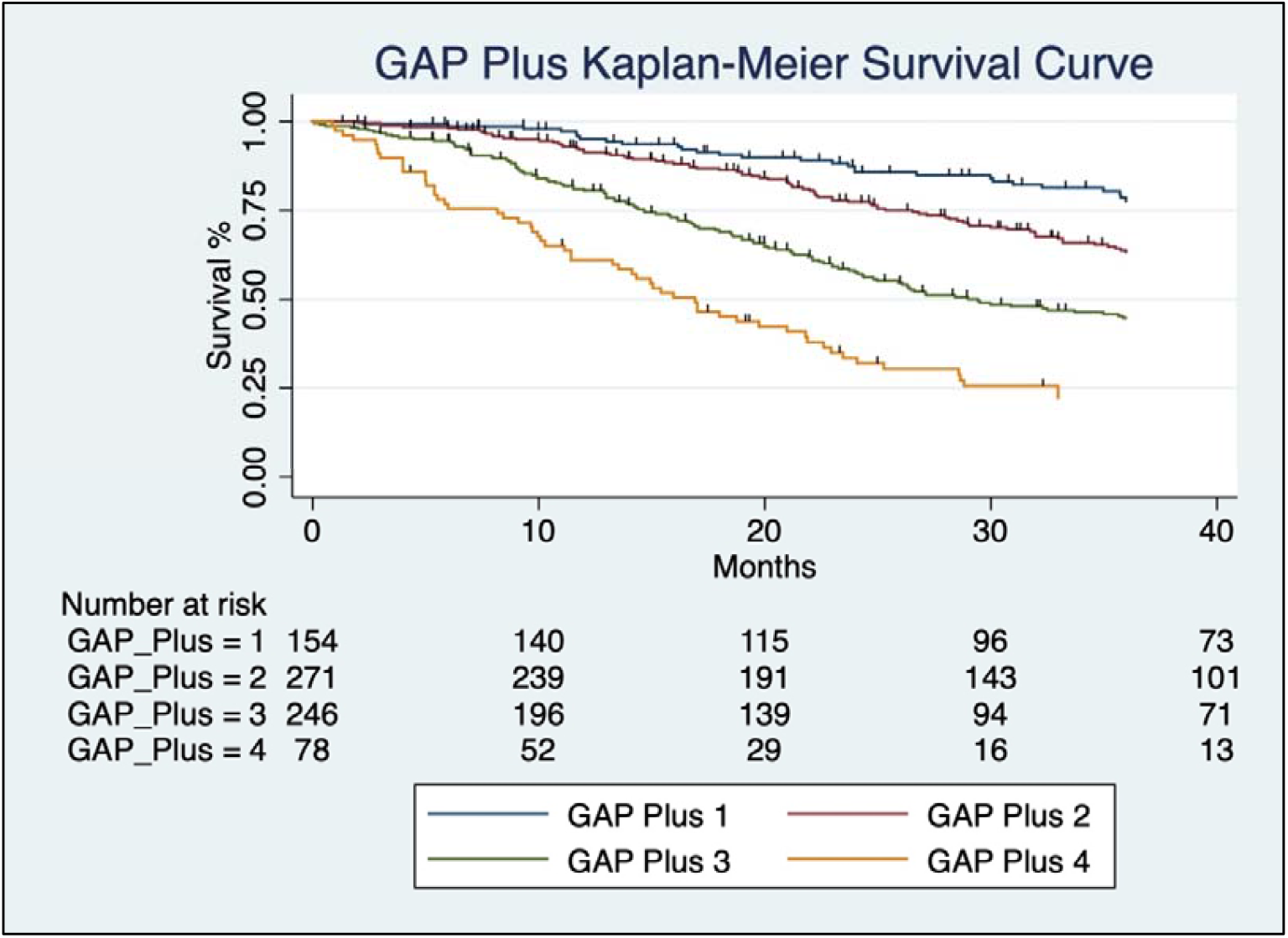
Kaplan-Meier survival curves for GAP-pus categories show all-cause mortality following a diagnosis of IPF with follow up extending to 40 months: A, All patients in combined cohort (n=999) were assigned to GAP-Plus Stages (initial GAP stage plus an additional 1 for patients with high, >/=2.9, NLR category at baseline): GAP-plus stage 1 (n=154), 2 (n=271) 3 (n=246) and 4 (n=78); The numbers of patients at risk at 10, 20, 30, 40 months for each of these groups is shown in the table immediately below the survival curves. The survival differences between the GAP-Plus groups of patients reached significance (HR 1.80, 95% CI 1.60-1.98; log rank test, p< 0.0001).

Univariate Cox proportional hazards models of the combined cohort data showed that patients in the high NLR category group had significantly higher mortality/progression to lung transplant when compared with patients in the low NLR group (HR 1.65, 95% CI 1.39-1.95; p<0·0001; not shown). NLR category remained significant when each site’s cohort was considered individually (Figure 4). Analysis was repeated in the combined cohort excluding all patients on known steroid therapy with a comparable result (HR 1.50, 95% CI 1.24-1.82; p < 0.0001; data not shown). Univariate regressions for GAP Index, GAP Stage and GAP-Plus Stage were all significant (p< 0.0001). Univariate regression of the individual GAP components (age, sex, FVC % predicted, TLco % predicted) was significant for all except sex. Cox regression for steroid use was also significant for transplant-free survival (p <0.0001). The analysis was repeated in the combined cohort excluding all patients on steroids and showed the same significances.

**Figure 4:**
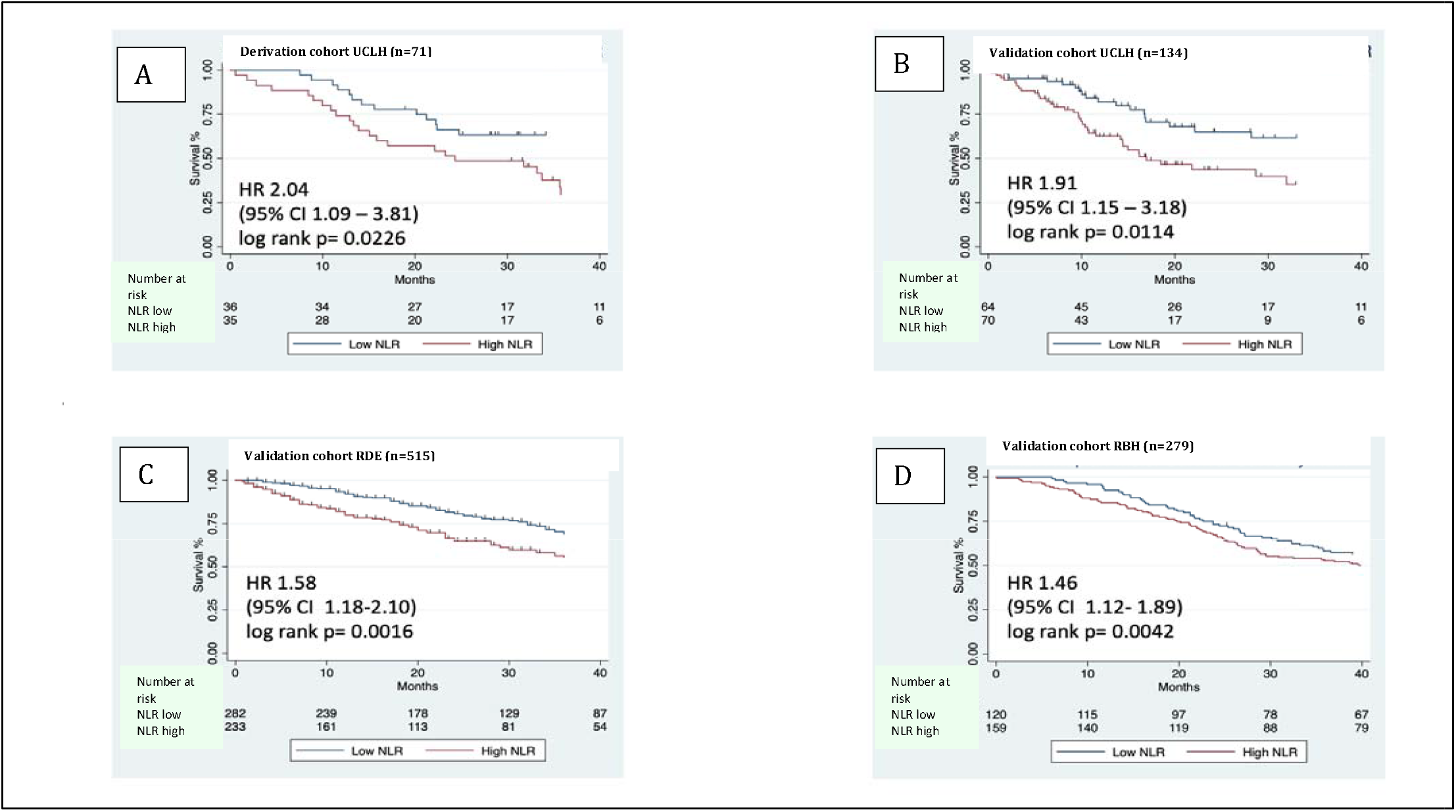
Kaplan Meir survival curves for NLR categories for cohorts shown for each centre show all-cause mortality following a diagnosis of IPF with follow up extending to 40 months. Graphs show patients i : A, derivation cohort, UCLH (n=71); B, validation cohort at UCLH (n=134); C, validation cohort RDE (n=515); D, Validation cohort RBH (n=279) who were assigned to low (<2.9) or high (>/=2.9) NLR category at baseline: The numbers of patients at risk at 10, 20, 30, 40 months for each of these groups is shown in the table immediately below the survival curves. HR, hazard ratio for event is shown for patients with high NLR compared to low NLR and include CI, confidence intervals, with P values for log rank tests shown on individual plots

Multivariate analysis showed that after adjusting for GAP Stage and use of steroids in the combined dataset a high NLR category was independently predictive of mortality/progression to lung transplant (HR 1.36, 95% CI 1.12-1.66; p=0.002). Repeating the analysis using the individual GAP components (age, sex, FVC%, TL_CO_%) as variables and again adjusting for steroids showed similar results (HR 1.26, 95% CI 1.03–1.55; p=0·027). Inputting NLR as a continuous variable was also independently predictive adjusted for the individual GAP components and steroids (HR 1.04, 95% CI 1.01-1.07; p=0.011) as well as when adjusted for GAP stage and steroid use (HR 1.05, 95% CI 1.02 -1.07, p= 0.001). All GAP components, except for sex, continued to be significant when adjusted for each other, NLR and steroid use.

Harrell’s concordance (C)-index prediction accuracy confirmed that the best performing prediction model was based on the component variables making up the GAP, with NLR as a continuous variable and adjusted for steroids. Incorporating NLR into GAP staging as GAP Plus and GAP Index Plus increased the model’s ability to predict patient mortality (C-index 0.673, 0.645-0.701, p<0.0001; Table 5).

**Table 5:**
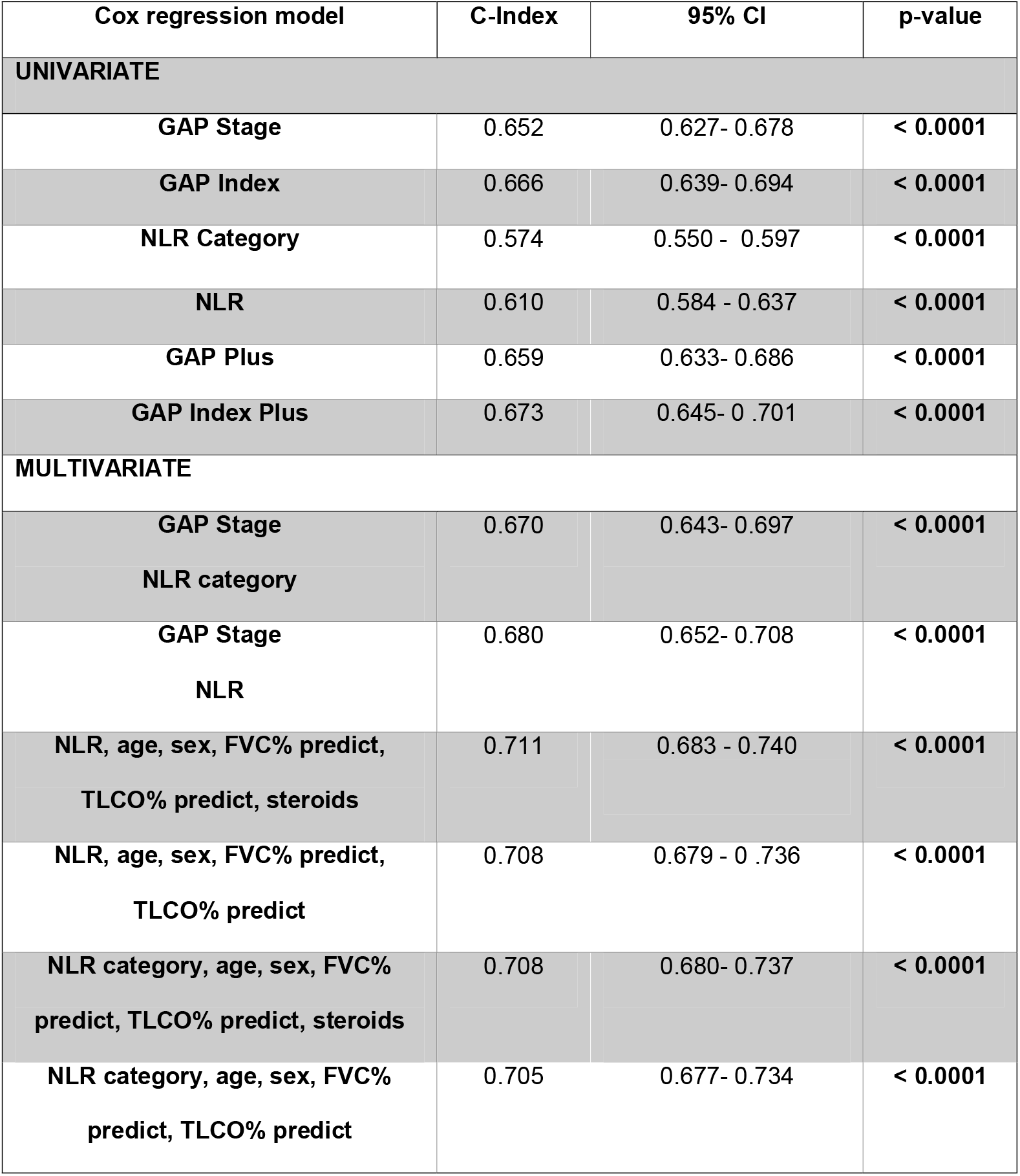
Harrell’s C-index performance in the pooled patient cohort (n=999) including steroid users in various predictive models.

Time-dependent ROC analysis in the pooled cohort for NLR demonstrated the continuous decline of the model’s predictive value with the passing of time. For example, AUC at 6 months is 0.728 which declines to an AUC of 0.598 at 48 months (Figure 5).

**Figure 5:**
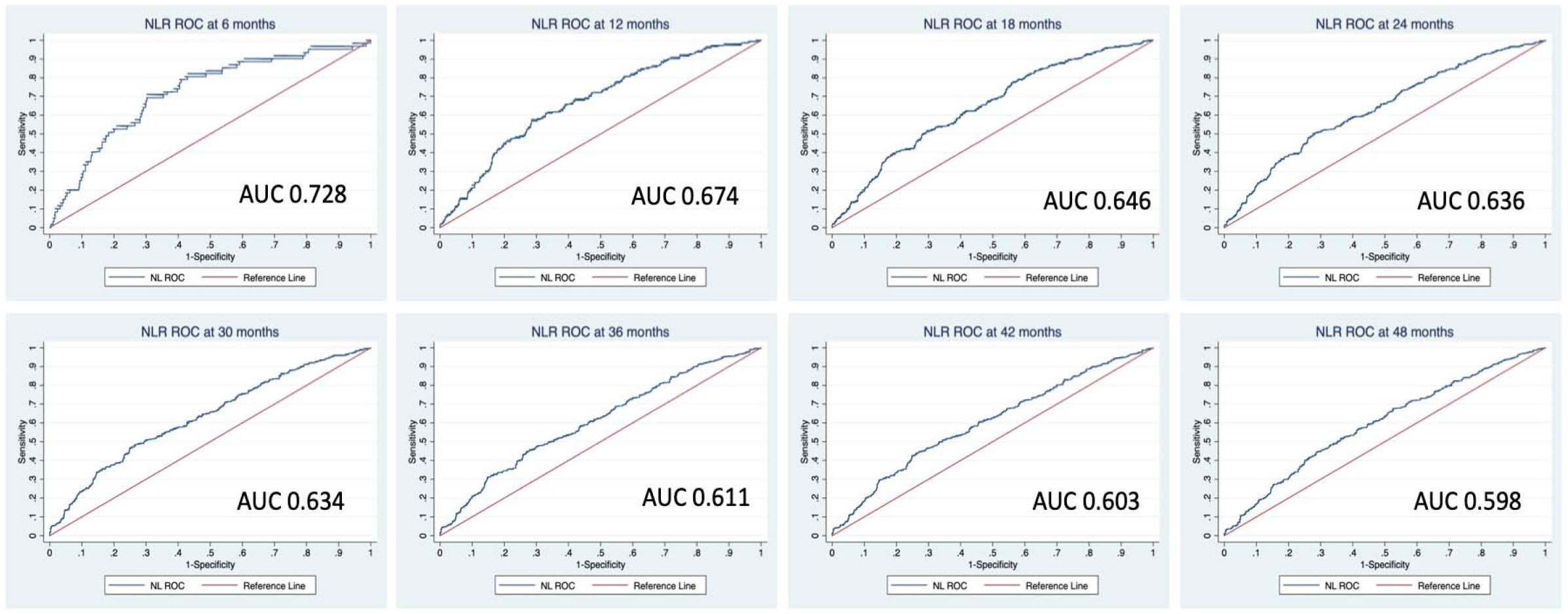
Ability of baseline NLR to predict all-cause mortality in patients with IPF decreases with time. Time dependent change of AUC and ROC in NLR at 6,12,18,24,30,36 and 42 months: AUC, area under the curve; ROC, receiver operator characteristic

## Discussion

IPF is a devastating disease with a variable clinical course. One of the most used prognostic cohort scoring systems is the GAP score. However, even within the same GAP stage, and particularly for moderately severe GAP stage II, patients may have very heterogenous outcomes. This has led to a concerted effort to identify better tools for individual patient risk stratification. The ideal biomarker would be measurable in the serum using a simple and widely available test and would predict prognosis and potentially response to therapy. Here we show that baseline NLR derived from a cheap and widely available routine blood test, identifies two classes of patients with IPF with significant differences in outcome. We go on to show that NLR can significantly refine the predictive capacity of the clinical GAP index.

The search for viable biomarkers has taken advantage of the rapidly expanding knowledge of IPF immunopathogenesis. Aberrant repair processes initiated by repetitive injury to the alveolar epithelium result in an exaggerated tissue remodelling response and fibrosis of the lung parenchyma. Proteins released from damaged epithelium and collagen degradation products can enter the systemic circulation, acting as markers of disease activity by proxy-the most promising of which include CA-19-9 ^18^, CA-125 ^18^ and CCL18 ^19^. Others that have been investigated include SP-D ^20^, MMP7 ^21^, osteopontin (OPN), periostin (PON), ICAM1 ^22^ and telomere length ^23^. In addition, neo-epitopes generated by the action of matrix metalloproteinases (MMPs) on collagen can be detected in the serum and Jenkins et al found that 6 of 12 of these were predictive for mortality ^24^. Other serum biomarkers include CD28, ICOS, LCK, ITK ^25^ alone or as part of a 52-gene RNA signature ^6^. More recently, attention has turned to imaging biomarkers including imaging quantification ^26^, measurements of glucose uptake in the lung with Positron Emission Tomography (PET) ^7^ and a combination of the two ^27^.

However, of these, only three biomarkers have been validated to refine the GAP staging system by identifying high and low risk patients within a given GAP stage. The 52-gene expression signature, an approach that requires calibration against a control cohort ^6^, a composite of OPN, PON, MMP-7 and ICAM-1^22^ and the Total-to-Background Ratio (TBR) calculated from 18F-FDG-PET imaging, with only the 52-gene expression validated in multiple independent cohorts.

NLR which is calculated from complete blood count with differential, is an inexpensive, easy to obtain, widely available and emerging marker of disease activity and prognosis in patients with chronic inflammatory diseases, cardiovascular diseases, and malignancies.

Previous studies have specifically considered NLR as a biomarker in IPF: Of two small single centre studies; the first ^1^ found NLR raised in 21 patients with IPF compared to 42 healthy controls but was not prognostic; the second study of 73 patients with IPF and 62 healthy controls found that NLR and monocyte lymphocyte ratio (MLR), but not platelet to lymphocyte ratio (PLR), associated with IPF and correlated negatively with FVC/ DLco ^2^. Another study measured NLR in bronchoalveolar lavage (BAL) samples from 59 patients with IPF and found that BAL NLR was inversely correlated with FVC measured at the same time as collection of the BAL sample ^3^. We presented our original discovery and validation cohort of 218 patients in 2018 ^17^prior to validation in external cohorts. Our initial findings were taken further by Nathan et al^4^, who included 1334 patients involved in ASCEND (Study 016; NCT01366209) and CAPACITY (Studies 004 and 006; NCT00287716 and NCT00287729) as a discovery cohort and placebo-treated IPF patients from two independent Phase III, trials of IFN-γ-1b (GIPF-001 (NCT00047645) and GIPF-007 (NCT00075998) as a validation cohort. Significant trends were observed between baseline NLR and PLR quartiles for various outcomes including: physiological decline; respiratory hospitalization; and all-cause mortality. However, the only consistent correlation in the discovery cohort was with baseline NLR and the composite endpoint of ‘absolute decline in 6MWD ≥50 m or death’ at 12 months, a finding that was not tested against a validatory cohort. Alongside this other groups were investigating circulating cellular biomarkers in IPF. Significant prognostic effects were found for monocyte count a finding validated in > 7000 patients with IPF from five independent cohorts ^28^and >2000 patients from a further four cohorts ^29^. However, the ability of monocyte count to enhance the predictive accuracy of GAP, although promising has not been validated ^30^in clinical cohorts.

In this retrospective study, we have extended the findings of Nathan et al. to investigate the role of NLR in multiple ‘real-life’ IPF cohorts with a longer follow-up period, to see if the current clinical prediction GAP score could be further refined. We analysed the NLR in a derivation cohort of patients and identified a median value of NLR that separated our discovery population into a high and low risk group for transplant-free survival with significant differences in mortality. We validated our findings in a prospective internal cohort of IPF patients and then in two external IPF cohorts provided by five other ILD specialist service centers in the UK. Furthermore, we showed, using a variety of statistical models, that the NLR is an independent risk factor for mortality, and addition of NLR risk profiles to the GAP index significantly increased the prediction accuracy of this clinical score.

Furthermore, we have been able to show that the addition of NLR data to GAP score refines the existing mortality prediction model by using C-index and ROC statistics. As expected, the more granular the data inputted the better the prediction model, hence the increased C-Index for a model using the individual components of the GAP Index rather than an overall score. This despite marked heterogeneity between the validation cohorts, with the RDE cohort being more recent (2011-2019) with a lower average GAP index, GAP stage and mean, and median NLR compared with the other cohorts. It is encouraging that NLR mortality prediction was robust despite this heterogeneity, pointing to wide-spread applicability

By using time-dependent ROC analysis we were able to calculate the decline in NLR’s predictive accuracy over time and establish that it is most accurate shortly after being measured, a time when indeed it is most useful. Many newly diagnosed patients are keen to discuss prognosis early and as clinicians we often refer to variable disease trajectory and the need to observe lung function over time to allow more accurate prognostication. However, these data suggest that even early mortality might be predictable from a high NLR at presentation and may expedite, for example, lung transplant assessment in appropriate patients. A similar decline in predictive accuracy with time has been shown for GAP and other biomarkers ^6,31^

The difference in median survival stratified by GAP stage was only significant for patients in the moderately severe GAP stage 2 and in those patients in whom the GAP stage could not be calculated. This probably reflects the small numbers of patients at GAP stage 1 and 3, although similar trends to significance in these groups are encouraging. In those patients in whom GAP could not be calculated it was interesting to observe that the overall median survival of 60 months was between the median survivals for GAP Stages 1 (73.7 months) and GAP Stage 2 (41 months). When the patients in GAP Stage 2 were stratified according to NLR category, a remarkable difference of survival became apparent on either side of this median with Gap Stage 2 and low NLR having a median survival of 83 months, and almost double that of those in GAP Stage 2 with a high NLR whose median survival was just 44 months. Refinement of this group of moderately severe IPF patients, GAP stage II, is particularly helpful as the outcomes of this group are the most heterogenous and it is here that clinical decision making can be most difficult.

GAP staging was not possible for those patients with incomplete lung function data, nearly always due to missing TLco readings. Gas transfer may be missing for several reasons, either the patient is unable to perform the test hence coded as a “3” (maximum) in the GAP index or a data quality issue. We found that patients with no TLco but in the low NLR group had a longer median survival than other patients in GAP stage 1, indicating that this subgroup may have been too well to warrant full lung function work up at the time of presentation. One additional feature of this study is our demonstration that NLR correlates with lung function, suggesting NLR may offer a cheap and quick screening test to fast-track high risk patients for early tertiary care review and urgent lung function. This might be especially relevant in current times of increased pressure and backlog on lung function testing due to the COVID-19 pandemic and in remote, and resource poor, areas where access to lung function is restricted.

It is unclear why NLR is raised in patients with IPF with decreased survival. We propose it could be a marker for ongoing inflammation. The term interstitial lung disease or ILD covers a group of over 200 different diseases with varying degrees of inflammation and/or fibrosis. It is unclear whether fibrosis is always preceded by inflammation, although this is more likely to be true for ILDs associated with underlying autoimmune rheumatic diseases. In such situations, NLR has been shown to predict development and extent of lung fibrosis, for example in systemic sclerosis, ^15^ and dermatomyositis/polymyositis ^16^. In this study, we demonstrate that NLR is also predictive in IPF, a disease in which inflammation is not thought to play a role, and indeed in which the use of immunosuppression in this disease has been shown to be harmful. It is unclear if NLR is alerting us to a potential role of inflammation in advancing interstitial inflammation or is highlighting a group of patients in which inflammation drives increased mortality from cardiovascular involvement. Disordered metabolism of carbohydrates, lipids, proteins, and hormones has been documented in lung, liver, and kidney fibrosis and metabolic dysregulation has been implicated in the pathogenesis of IPF ^32^, potentially offering a new target for fibrosis therapy.

The predictive ability of NLR may hint more directly at a role for neutrophilic inflammation in the pathogenesis of IPF. We have known for a long time that the percentage of neutrophils in the BAL of patients with IPF correlates with a poor outcome ^33^. Molyneaux et al. have shown that BAL neutrophilia is associated with both increased microbiome burden and progressive IPF ^34^, with subtle changes in the microbiome implicated in the initiation and progression of IPF in the absence of identified infection ^35^. The increased bacterial burden of IPF appears to be in the airway, proximal to the actual fibrotic remodeling of the parenchyma, with very low levels of bacteria identified in IPF parenchymal lung tissue ^36^. However, such changes are unlikely to cause increases in systemic neutrophilia and NLR in the absence of overt infection. In our study, we excluded patients diagnosed clinically with infection and started on antibiotics, and those in whom the C reactive protein (CRP) was greater than 20 mg/L.

If we do not think NLR is detecting occult infection, then why is it such a powerful marker? One developing line of enquiry is that the lung is responsible not just for gas-exchange but also plays a crucial role in leukocyte homeostasis. There is increasing evidence that the lung may orchestrate the disposal of aged neutrophils, by targeting them for recirculation to, and disposal in, the bone marrow. In a mouse model the inability of the lung to clear aged neutrophils resulted in a pulmonary fibrosis ^37^. As well as neutrophil activation, other groups have noted phenotypic changes in circulating leukocytes, for example CD28 downregulation on CD4 cells, perhaps reflecting T cell exhaustion, and 4 T cell genes (CD24, ICOS, LCK and ITK) are part of the 52 gene signature that is associated with a poor disease outcome ^6^. Interestingly we found that the neutrophil count is not as strong a predictor of mortality in IPF as NLR, suggesting that both neutrophil activation and lymphocyte exhaustion may be relevant.

Despite the reproducibility of our findings there are some caveats. We did not determine the specificity of NLR to IPF as opposed to other ILDs. However, we have previously reported that within the ILD cohort from RDE/NB/TS/UHL although high baseline NLR predict outcomes in IPF this was not the case in patients with chronic hypersensitivity pneumonitis ^5^. Secondly, the majority of the patients were in the pre-anti fibrotic era and were treated with corticosteroids. However, we have shown that use of corticosteroids did not influence the validity of NLR. We have only limited longitudinal data, and there is a suggestion that patients will change their profiles but how this relates to their prognosis is not clear. Nathan et al found NLR change may be an even more robust prognostic biomarker than baseline NLR but may be less suitable as a predictive biomarker for patients receiving treatment with antifibrotics ^4^. The main limitation of this retrospective study is linked to missing, and at times, poor quality data. Our data does offer impetus to the idea that NLR should be evaluated as part of a prospective clinical trial as a secondary or an exploratory endpoint.

In summary, we have demonstrated and validated that NLR, an easily, widely available, cheap and reproducible test, is an independent prognostic biomarker that can be evaluated at diagnosis in patients with IPF and may inform future management of these patients. There is an enhanced cohort outcome prediction accuracy when NLR is added to GAP score suggesting that NLR may be useful not only as a stratification marker, but also a predictor for disease monitoring in IPF. One striking observation is that NLR correlates with lung function (FVC and DLco) and may be particularly helpful in assessing clinical priorities in situations where lung function is not easily available, such as in remote areas and during the pandemic, or cannot be performed by the patient. Further evaluation of the utility of NLR measurement for therapeutic decision making is warranted.

## Data Availability

All data produced in the present study are available upon reasonable request to the authors

## Data sharing statement

Data collected for the study may be accessed after approval of a proposal and with a signed data access agreement with the individual investigators that manage the patient databases.

## Ethical Approval

Ethical approval (and any ensuing amendments) was granted by the HRA and Health and Care Research Wales (HCRW). (REC reference: 18/LO/0937) (appendix x). Site specific and local R&D approvals were granted by each participating hospital site. The study was sponsored by University College London (UCL).

## Supplementary Table

**Table S1:**
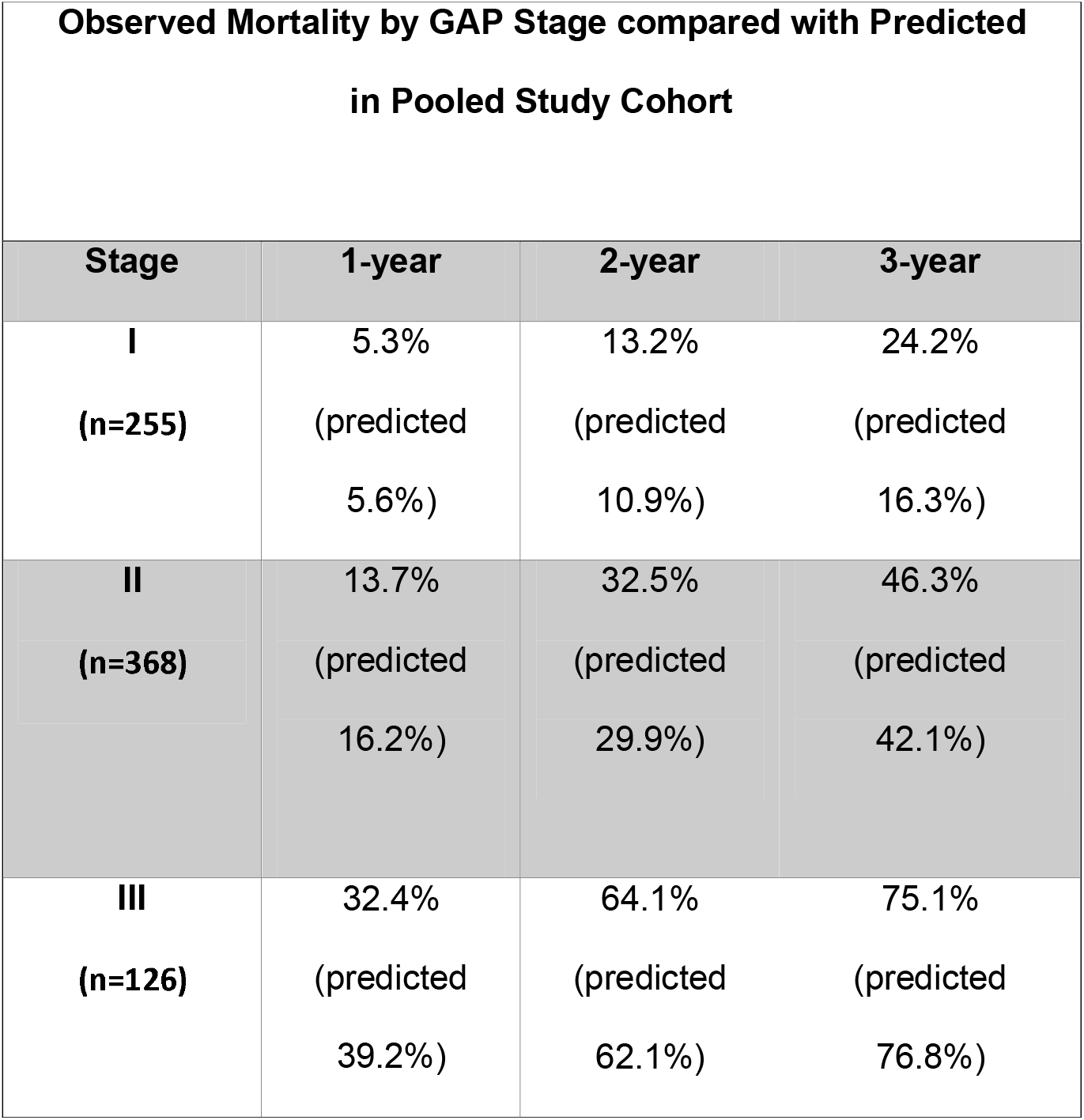
Observed (and Predicted) Mortality by GAP Stage for our cohort (n=999) compared to the literature predicted values.

STROBE Statement—Checklist of items that should be included in reports of ***cohort studies***

|  | Item No | Recommendation | Page No |
| --- | --- | --- | --- |
| **Title and abstract** | 1 | (a) Indicate the study's design with a commonly used term in the title or the abstract<br>(b) Provide in the abstract an informative and balanced summary of what was done and what was found | 1 |
| **Introduction** |  |  |  |
| Background/rationale | 2 | Explain the scientific background and rationale for the investigation being reported | 2 |
| Objectives | 3 | State specific objectives, including any prespecified hypotheses | 2 |
| **Methods** |  |  |  |
| Study design | 4 | Present key elements of study design early in the paper | 2 |
| Setting | 5 | Describe the setting, locations, and relevant dates, including periods of recruitment, exposure, follow-up, and data collection | 3 |
| Participants | 6 | (a) Give the eligibility criteria, and the sources and methods of selection of participants. Describe methods of follow-up<br>(b) For matched studies, give matching criteria and number of exposed and unexposed | 3 |
| Variables | 7 | Clearly define all outcomes, exposures, predictors, potential confounders, and effect modifiers. Give diagnostic criteria, if applicable | 3 |
| Data sources/measurement | 8* | For each variable of interest, give sources of data and details of methods of assessment (measurement). Describe comparability of assessment methods if there is more than one group | 3 |
| Bias | 9 | Describe any efforts to address potential sources of bias | 6 |
| Study size | 10 | Explain how the study size was arrived at | 3 |
| Quantitative variables | 11 | Explain how quantitative variables were handled in the analyses. If applicable, describe which groupings were chosen and why | 2 |
| Statistical methods | 12 | (a) Describe all statistical methods, including those used to control for confounding<br>(b) Describe any methods used to examine subgroups and interactions<br>(c) Explain how missing data were addressed<br>(d) If applicable, explain how loss to follow-up was addressed<br>(e) Describe any sensitivity analyses | 3 and 5 |
| **Results** |  |  |  |
| Participants | 13* | (a) Report numbers of individuals at each stage of study—eg numbers potentially eligible, examined for eligibility, confirmed eligible, included in the study, completing follow-up, and analysed<br>(b) Give reasons for non-participation at each stage<br>(c) Consider use of a flow diagram | 5 |
| Descriptive data | 14* | (a) Give characteristics of study participants (eg demographic, clinical, social) and information on exposures and potential confounders<br>(b) Indicate number of participants with missing data for each variable of interest<br>(c) Summarise follow-up time (eg, average and total amount) | 15 |
| Outcome data | 15* | Report numbers of outcome events or summary measures over time |  |
| Main results | 16 | (a) Give unadjusted estimates and, if applicable, confounder-adjusted estimates and their precision (eg, 95% confidence interval). Make clear which confounders were adjusted for and why they were included<br>(b) Report category boundaries when continuous variables were categorized<br>(c) If relevant, consider translating estimates of relative risk into absolute risk for a meaningful time period | 17 |
| Other analyses | 17 | Report other analyses done—eg analyses of subgroups and interactions, and sensitivity analyses | 17 |
| **Discussion** |  |  |  |
| Key results | 18 | Summarise key results with reference to study objectives | 8 |
| Limitations | 19 | Discuss limitations of the study, taking into account sources of potential bias or imprecision. Discuss both direction and magnitude of any potential bias | 10 |
| Interpretation | 20 | Give a cautious overall interpretation of results considering objectives, limitations, multiplicity of analyses, results from similar studies, and other relevant evidence | 9 |
| Generalisability | 21 | Discuss the generalisability (external validity) of the study results | 10 |
| **Other information** |  |  |  |
| Funding | 22 | Give the source of funding and the role of the funders for the present study and, if applicable, for the original study on which the present article is based | 11 |

\*Give information separately for exposed and unexposed groups.
**Note:** An Explanation and Elaboration article discusses each checklist item and gives methodological background and published examples of transparent reporting. The STROBE checklist is best used in conjunction with this article (freely available on the Web sites of PLoS Medicine at http://www.plosmedicine.org/, Annals of Internal Medicine at http://www.annals.org/, and Epidemiology at http://www.epidem.com/). Information on the STROBE Initiative is available at http://www.strobe-statement.org.

